# Validation and context-dependent effects of a prostate cancer polygenic risk score in the All of Us Research Program

**DOI:** 10.1101/2025.10.01.25337107

**Authors:** Shuyan Cheng, Austin Hammermeister Suger, Louisa B. Goss, Jiachen Zhang, Harriett Fuller, Boya Guo, Sara Lindström, Burcu F. Darst

**Affiliations:** School of Public Health, University of Washington, Seattle, WA, USA; Division of Public Health Sciences, Fred Hutchinson Cancer Center, Seattle, WA, USA

**Keywords:** Polygenic risk score, prostate cancer, context-dependent effects, large-scale biobanks, phenome-wide association study, risk modeling

## Abstract

Polygenic risk scores (PRSs) have demonstrated strong potential for improving prostate cancer risk stratification. However, it is unknown whether the clinical utility of prostate cancer PRS vary by demographic, lifestyle, and socioeconomic factors. We validated a previously developed multi-ancestry PRS of 451 prostate cancer risk variants and evaluated context-dependent effects using genetic and clinical data from the diverse All of Us Research Program, including 7,577 cases and 90,608 controls across six genetic ancestry groups. In ancestry-stratified testing, the PRS showed strong associations with prostate cancer risk, with odds ratios (ORs) per standard deviation (SD) increase ranging from 1.61 (95% CI=1.02-2.64, P=0.05) in Middle Eastern to 2.19 (95% CI=1.98-2.42, P=2.2×10^-51^) in American populations. Age-stratified analyses showed an overall reduced PRS effect with increasing age. Across modifiable lifestyle and healthcare access factors, PRS effects were larger in those with higher body mass index (OR ranging from 1.71-2.17 in underweight to obese individuals, P=0.02), in never or former smokers vs. current smokers (OR=2.06, 2.37, and 1.93, respectively, P=0.06), and in those recently accessing healthcare (OR=2.21 vs. 1.88, P=0.05), highlighting important context-specific modifiers. We did not observe context-dependent effects by other socioeconomic factors, such as income, education, and insurance. In a phenome-wide association study (PheWAS), the PRS was associated with 14 clinical outcomes, including known prostate cancer-related conditions. These findings confirm the predictive strength of the multi-ancestry prostate cancer PRS across diverse populations and underscore the importance of accounting for demographic, lifestyle, and healthcare-related contexts when applying PRS in clinical and public health settings.

## Main text

Prostate cancer is one of the most commonly diagnosed cancers among men worldwide and a leading cause of cancer-related mortality^1^. Polygenic risk scores (PRS), which aggregate the effects of multiple genetic variants, have shown strong potential in stratifying prostate cancer risk and guiding personalized prevention and early detection strategies^2–5^. However, the clinical utility of prostate cancer PRS may vary across populations due to differences in demographic, lifestyle, and socioeconomic factors^6–9^. Healthy lifestyle factors—such as regular physical activity^10–12^, non-smoking^13^, and lower adiposity^14–17^—have been associated with reduced risk of lethal prostate cancer, though most evidence to date is limited to European descent populations. Understanding whether prostate cancer PRS have context-dependent effects in multi-ancestry populations is essential to ensure equitable and effective implementation of PRS-informed care. The All of Us (AoU) Research Program, with its large, diverse participant base and rich genomic and electronic health record data, provides an ideal platform to validate prostate cancer PRS in a multi-ancestry population-based cohort and explore how its predictive performance varies across different sociodemographic and population contexts^18,19^.

In this study, we leveraged AoU to validate a previously developed multi-ancestry prostate cancer PRS of 451 risk variants^2,3^ and examine its performance across diverse populations and clinical contexts to inform equitable precision health strategies (**Supplemental Material**). Participants included 7,577 prostate cancer cases and 90,608 male sex controls from AoU who were genetically similar to European (EUR, 5,757 cases and 54,751 controls), African (AFR, 1,212 cases and 20,193 controls), American (AMR, 492 cases and 12,807 controls), East Asian (EAS, 63 cases and 1,512 controls), South Asian (SAS, 27 cases and 1,017 controls), and Middle Eastern (MID, 26 cases and 328 controls) ancestry populations. We observed strong associations between PRS and prostate cancer risk across all genetic ancestry groups. The largest effect was observed in individuals of AMR ancestry (OR = 2.19 per SD unit increase in PRS, 95% CI = 1.98– 2.42, P = 2.2×10^-51^), followed closely by EUR (OR = 2.13, 95% CI = 2.07–2.20, P < 2.2×10^-308^), EAS (OR = 2.12, 95% CI = 1.60–2.85, P = 3.0×10^-07^), AFR (OR = 1.99, 95% CI = 1.87–2.13, P = 1.2×10^-96^), SAS (OR = 1.69, 95% CI = 1.14–2.53, P = 0.009), and MID (OR = 1.61, 95% CI = 1.02–2.64, P = 0.05) ancestry populations (**Figure 1** and **Supplemental Table 1**). Comparing individuals in the top PRS decile (90–100%) to those with average genetic risk (40–60%), prostate cancer OR were 4.67 (95% CI = 3.39–6.50, P = 1.9×10^-20^) for AMR, 4.39 (95% CI = 3.99–4.83, P = 1.8×10^-203^) for EUR, and 3.71 (95% CI = 3.02–4.58, P = 9.8×10^-35^) for AFR ancestry populations (**Figure 1** and **Supplemental Table 1**; sample sizes were too small to investigate PRS deciles in EAS, SAS, and MID). Across all populations, we observed a +0.05 improvement in the area under the curve (AUC) when adding the PRS to a model of age and 10 principal components (AUC=0.82, 95% CI=0.81–0.82 vs AUC=0.77, 95% CI=0.77–0.78, DeLong test P-value = 4.6×10^-160^), with similar AUC improvements observed for most ancestry populations (**Supplemental Figure 1**).

**Figure 1.**
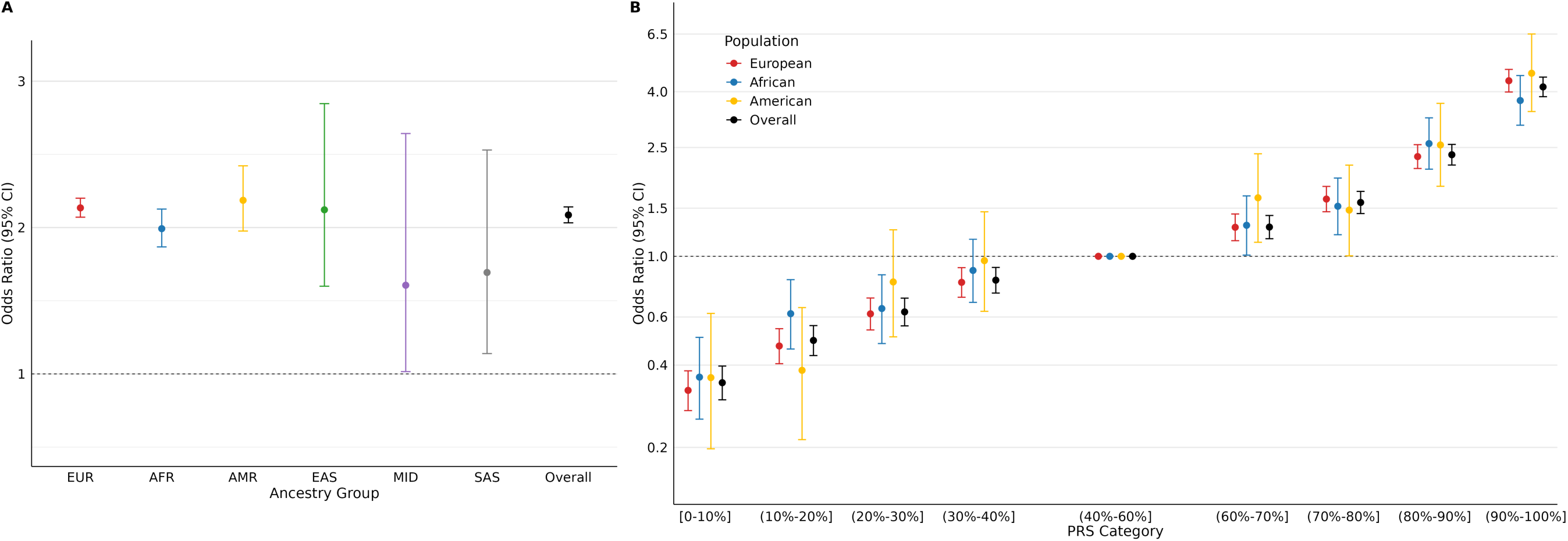
Association between PRS and prostate cancer risk across AoU genetic ancestry populations. A) Association results using the continuous standardized PRS. B) Association results using PRS decile categories. Note that population-specific results for PRS deciles are limited to AFR, AMR, and EUR ancestry populations due to the limited number of cases in other ancestry populations, while the overall results include all populations.

Consistent with our previous findings^3,4^, PRS associations were strongest among younger individuals and declined with age at sample draw in AFR (OR = 2.41, 95% CI = 2.00–2.91 for those ≤55 years; OR = 2.06, 95% CI = 1.90–2.24 for those 55–70 years; and OR = 1.59, 95% CI = 1.40–1.81 for those >70 years; heterogeneity P = 3.1×10^-4^) and AMR (OR = 3.29, 95% CI = 2.25–4.87 for those ≤55 years; OR = 2.20, 95% CI = 1.91–2.53 for those 55–70 years; and OR = 1.93, 95% CI = 1.65–2.27 for those >70 years; heterogeneity P = 0.04) ancestry populations (**Figure 2, Supplemental Table 2**, and **Supplemental Material**). Among EUR ancestry individuals, the effect of PRS was strongest in the 55–70 age group (OR = 2.21, 95% CI = 2.10–2.32) and did not significantly differ by age (heterogeneity P = 0.09), which could potentially reflect misclassification of undiagnosed prostate cancer cases in the younger age group. Indeed, in our AoU sample, EUR ancestry individuals were diagnosed at a significantly older age (mean = 67.40 years, SE = 0.11) compared to AFR (mean = 62.88 years, SE = 0.23; P = 1.9×10^-62^) and AMR (mean = 65.21 years, SE = 0.38, P = 4.6×10^-08^) ancestry individuals.

**Figure 2.**
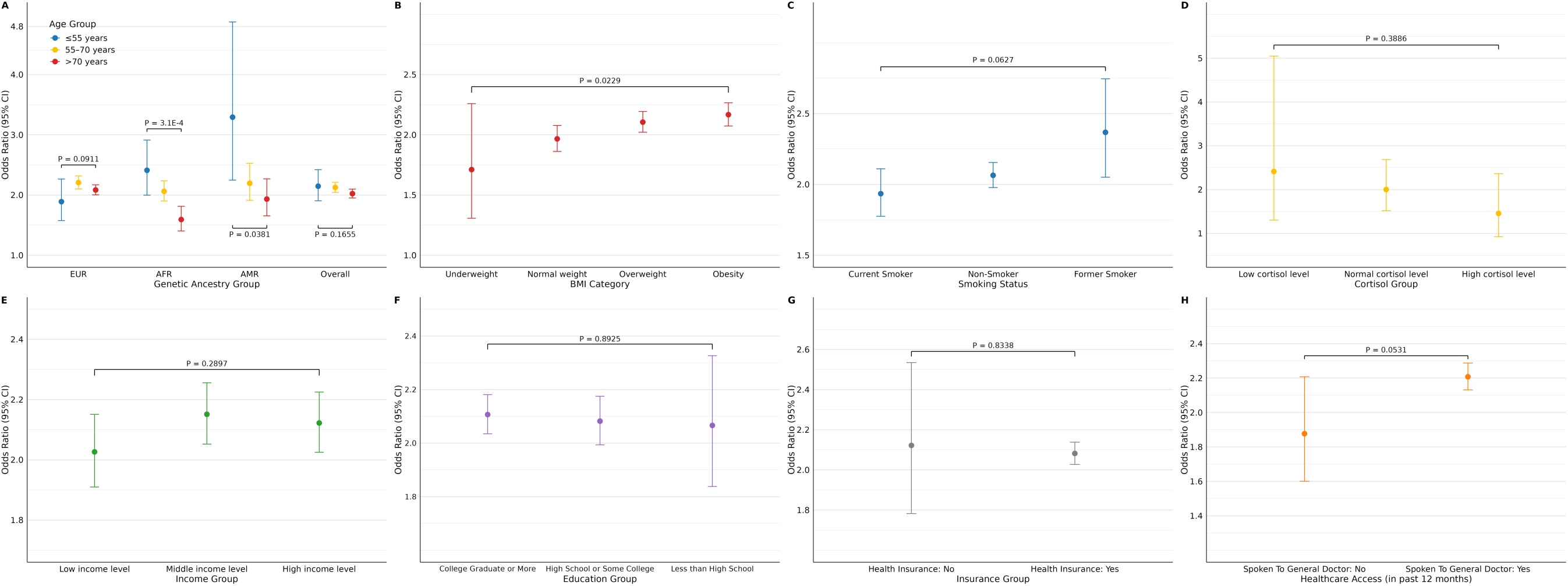
Association between PRS and prostate cancer risk stratified by demographic, lifestyle, and social contexts, including (A) age at sample draw, (B) BMI, (C) smoking status, (D) cortisol levels, (E) income, (F) education, (G) insurance status, and (H) healthcare access. OR with 95% CI are reported, with heterogeneity P-values from Cochrane’s Q-tests. Panel A is shown by EUR, AFR, and AMR ancestry group (EAS, MID, and SAS sample sizes were too limited to investigate stratified by age); other factors were not evaluated by ancestry due to limited sample sizes.

To account for different ways of defining age, we also performed analyses using age at diagnosis for cases and current age for controls, as is often done in case-control studies. In EUR ancestry, the association was largest in the ≤55 age group (OR = 2.22, 95% CI = 1.99-2.47) and declined with age (55–70 age group OR = 2.07, 95% CI = 1.98-2.16 and >70 age group OR = 1.88, 95% CI = 1.79-1.97, heterogeneity P = 0.002; **Supplemental Table 2**). However, this approach may not properly account for age differences between cases and controls. Re-evaluating age-stratified PRS associations as the AoU cohort ages will be important to confirm our findings. We also validated our previous finding that higher PRS was associated with a younger age at diagnosis^3^. Across all populations, we observed a 64.70 (0.31) mean (SE) age at diagnosis for those in the 90-100% PRS compared to 67.80 (0.34) for those in the 0-10% PRS category (P = 3.1×10^-11^), with consistent findings within each population (**Supplemental Figure 2** and **Supplemental Table 3**). This supports the interpretation that PRS has greater predictive value for earlier-onset prostate cancer across ancestries.

We further assessed whether the PRS had context-dependent associations with prostate cancer risk that varied by key lifestyle factors among all populations (**Figure 2, Supplemental Table 2, and Supplemental Material**). We observed larger PRS effects with higher BMI: ORs were 1.71 (95% CI = 1.31–2.26, P = 1.1×10^-04^) for underweight, 1.97 (95% CI = 1.86–2.08, P = 2.7×10^-129^) for normal weight, 2.11 (95% CI = 2.02–2.19, P = 3.6×10^-280^) for overweight, and 2.17 (95% CI = 2.07–2.27, P = 1.3×10^-255^) for individuals with obesity (heterogeneity P = 0.02). We observed nominal (P<0.1) evidence that PRS performance varied by smoking status: the OR was highest in former smokers (OR = 2.37, 95% CI = 2.05–2.75, P = 4.1×10^-31^), followed by non-smokers (OR = 2.06, 95% CI = 1.98–2.15, P = 7.1×10^-242^) and current smokers (OR = 1.93, 95% CI = 1.78–2.11, P = 1.3×10^-50^; heterogeneity P = 0.06). We also investigated differences by cortisol levels, which are typically lower in individuals who maintain regular exercise, adequate sleep, and a healthy diet^20,21^. Our findings suggest that PRS associations were strongest in men with low morning serum cortisol levels (OR = 2.41, 95% CI = 1.30–5.05, P = 9.7×10^-03^), followed by those with normal (OR = 2.00, 95% CI = 1.52–2.69, P = 1.8×10^-06^) and high (OR = 1.45, 95% CI = 0.92–2.36, P = 0.11) cortisol levels; however, these differences were not significant (heterogeneity P = 0.39), possibly due to the limited number of participants with morning cortisol measurements (118 cases and 892 controls), and warrant further investigation. Collectively, these results suggest that the effect of PRS on prostate cancer risk may vary by lifestyle-related factors.

To evaluate PRS performance across social contexts, we examined associations stratified by income, education, insurance status, and healthcare access across all populations (**Supplemental Material**). The effect of PRS on prostate cancer risk was consistent across income levels (heterogeneity P = 0.29), education groups (heterogeneity P = 0.89), and health insurance status (heterogeneity P = 0.83; **Figure 2** and **Supplemental Table 2**), suggesting that these socioeconomic factors may not impact the performance of prostate cancer PRS. Despite the lack of effect heterogeneity evidence, we did observe evidence of a larger PRS effect when comparing individuals with middle (OR = 2.15, 95% CI = 2.05–2.26, P = 4.1×10^-221^) vs low income (OR = 2.03, 95% CI = 1.91–2.15, P = 1.0×10^-119^; interaction P = 0.01); however, this difference is likely not clinically significant. Individuals who reported speaking to a general doctor in the past year showed a stronger association (OR = 2.21, 95% CI = 2.13–2.29, P < 2.2×10^-308^) compared to those who had not (OR = 1.88, 95% CI = 1.60–2.21, P = 1.6×10^-14^; heterogeneity P = 0.05), which likely reflects that healthcare access or engagement influences the likelihood of being properly diagnosed with prostate cancer.

A phenome-wide association study (PheWAS) was conducted to explore potential clinical consequences of having an elevated prostate cancer PRS. Among male sex participants, the PRS showed significant positive associations with 14 phenotypes, including conditions commonly related to prostate cancer diagnosis, treatment, and progression. These included elevated prostate-specific antigen (PSA), radiation-related effects, urinary incontinence, erectile dysfunction, hematuria, and secondary malignancy of bone—the most common site of metastasis for prostate cancer^22^. Several additional urinary and prostate-related disorders, such as inflammatory diseases of the prostate, urinary tract infection, and bladder neck obstruction were also identified (**Figure 3; Supplementary Table 4**). Findings were consistent but weaker in a separate PheWAS across sexes, with a noted lack of association with breast and other cancers, consistent with previous findings^23–25^ (**Supplementary Table 5**). These findings suggest that the prostate cancer PRS is fairly specific to prostate cancer and prostate cancer-related conditions.

**Figure 3.**
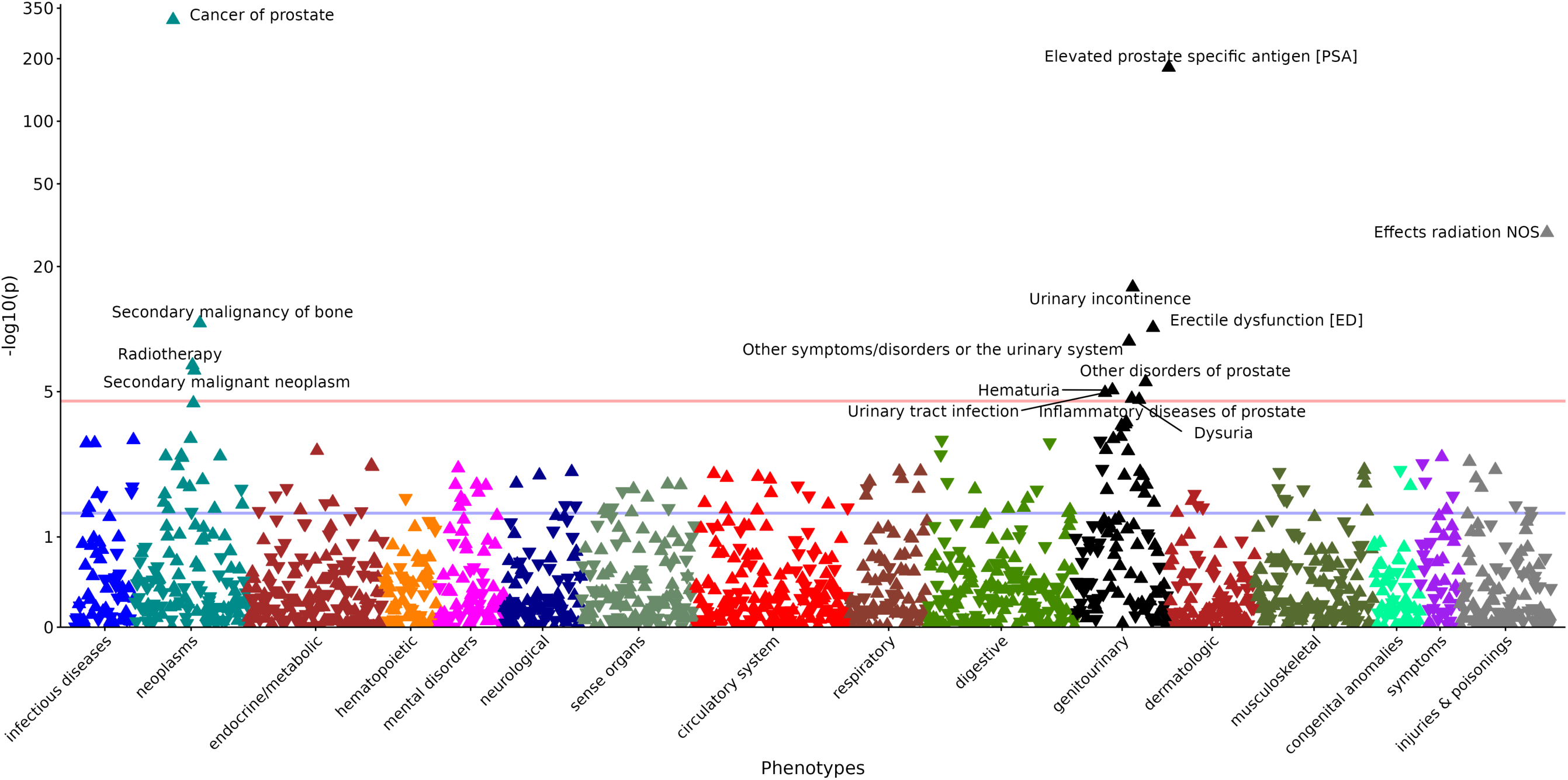
Phenome-wide association study of prostate cancer PRS in male sex participants.

Findings from our investigation support the predictive validity of the prostate cancer PRS across genetic ancestry populations and provide novel evidence of PRS validation in EAS, MID, and SAS ancestry populations, although the limited sample sizes of these populations warrant further validation. Our findings also offer insights into the contextual factors influencing genetic risk, with evidence suggesting that the PRS could be a better predictor of prostate cancer risk among those with a healthy lifestyle, including individuals who do not currently smoke. This corroborates evidence of the PRS having a stronger effect in younger individuals reported here and elsewhere for prostate cancer and other traits^4,26,27^, suggesting that the contribution of genetics to prostate cancer risk is greater in individuals who have not accumulated many environmental risk factors. However, contrary to these findings, the strength of PRS associations increased with higher BMI. Prior research has shown that increased BMI is inversely associated with prostate cancer risk, which could be attributed to higher BMI being associated with lower PSA and challenges to detect an abnormal finding during a digital rectal exam^14,28^. These both can reduce the sensitivity of screening, leading to detection bias and delayed diagnosis. Accordingly, studies have found that increased total adiposity (measured by BMI and visceral fat) and central adiposity (measured by waist circumference) are associated with increased risk of advanced and lethal prostate cancer^15–17^. In this context, PRS may appear more predictive in individuals with higher BMI because only those with a stronger genetic predisposition or clinically significant prostate cancer are likely to be detected through less effective screenings. The absence of tumor stage, grade, and prostate cancer-specific mortality in AoU limited our ability to further investigate this relationship. However, our findings suggest that among higher BMI individuals, the prostate cancer PRS is highly effective and could provide useful additional information to guide risk-based screening strategies and mitigate the detection bias in this group.

Our findings provide a better understanding of the contexts that impact the ability of PRS to predict prostate cancer risk, underscoring the importance of accounting for demographic, lifestyle, and healthcare-related contexts when implementing and interpreting PRS in clinical and public health settings. It will be important for future research to consider additional environmental, behavioral, and social factors as well as advanced and lethal prostate cancer outcomes. For instance, previous studies have found that exposure to agricultural pesticides, neighborhood factors, and dietary factors are associated with lethal prostate cancer risk^29–33^. Such investigations may be facilitated by AoU as additional survey responses, electronic health records, and geocoded data become available and would be further strengthened by the inclusion of cancer stage, grade, and outcomes information, which remains a critical gap for advancing this line of research.

## Data Availability

All data produced in the present study are available upon reasonable request to the authors

## Acknowledgements

This work was supported by the National Cancer Institute at the National Institutes of Health (R00 CA246063, R03 CA287235, U01 CA261339, Pacific Northwest Prostate Cancer SPORE P50 CA097186, Cancer Center Support Grant P30 CA015704), The Fred Hutch Translational Data Science New Collaboration Award, an award from the Andy Hill Cancer Research Endowment Distinguished Researchers Program, a Fred Hutch/University of Washington SPORE Career Enhancement Program award, the Prostate Cancer Foundation (21YOUN11), and the Institute for Prostate Cancer Research. We gratefully acknowledge *All of Us* participants for their contributions, without whom this research would not have been possible. We also thank the National Institutes of Health’s *All of Us* Research Program for making available the participant data examined in this study.

## Author contributions

Study conception/design: S.C., B.F.D.; data analysis: S.C., A.H.S., B.G., B.F.D.; interpretation of data: S.C., H.F., B.G., B.F.D.; drafted the manuscript: S.C., B.F.D.; contributed to manuscript revisions: S.C., A.H.S., L.B.G., J.Z., H.F., B.G., S.L., B.F.D.; all authors approved of the final manuscript.

## Declaration of interests

The authors declare no competing interests.

## Web resources

PGS catalogue: https://www.pgscatalog.org/

*All of Us* Research Program: https://allofus.nih.gov/

*All of Us* Researcher Workbench: https://workbench.researchallofus.org/login

## Data and code availability

This study used data from the *All of Us* Research Program’s Controlled Tier Dataset version 8, available to authorized users on the Researcher Workbench. The prostate cancer PRS of 451 variants is available on the PGS Catalog under accession numbers PGS003765 (for build 37 coordinates; https://www.pgscatalog.org/score/PGS003765/) and PGS003766 (for build 38 coordinates; https://www.pgscatalog.org/score/PGS003766/).

## Supplemental Material

### Participants

This study utilized data from the AoU Research Program Controlled Tier version 8 dataset^19^. Participants were included if their genetic sex was male and they had available genotype and EHR data. Prostate cancer was defined using a modified version of the eMERGE prostate cancer phenotyping algorithm^34^, where cases had at least one prostate cancer-related diagnostic code (International Classification of Diseases (ICD) 9 codes 185.* or V10.46, ICD-10 codes C61.*, Z85.46, or R97.21, or corresponding SNOMED concepts in AoU for malignant neoplasm of the prostate), with at least two distinct dates of occurrence. Controls were defined as male participants who had no evidence of prostate cancer based on ICD diagnostic codes, SNOMED terms, prostatectomy based on ICD or Current Procedural Terminology codes, or self-reported prostate cancer history. To reduce misclassification, controls were over 40 years of age at their most recent clinical encounter. The final analytic sample comprised 7,577 prostate cancer cases and 90,608 controls across six genetic ancestry groups (described below). All included individuals had complete covariate information, including age at sample draw and the first 10 principal components of ancestry, and passed standard quality control thresholds^35^.

All participants provided informed consent through the program’s eConsent process, which included primary consent, HIPAA authorization for research use of electronic health records and other external data, and genomic result return consent, as required by the AoU protocol. This study was approved by the Fred Hutchinson Cancer Center Institutional Review Board.

### Population Descriptors

We evaluated PRS performance separately by genetic ancestry groups. As previously described^35^, genetic ancestry was inferred within AoU using a harmonized pipeline trained on variants from the Human Genome Diversity Project (HGDP) and 1000 Genomes Project reference samples. A random forest classifier was applied to the first 16 principal components of each participant’s genotypes to estimate genetic similarity to global reference populations. Individuals were assigned to one of six categories based on the gnomAD, HGDP, and 1000 Genomes labels: African (AFR), American (AMR), East Asian (EAS), European (EUR), Middle Eastern (MID), or South Asian (SAS). Individuals are assigned to the most probable ancestry group based on classification confidence scores.

### Context-Dependent Variables

To examine context-dependent effects of the PRS, stratified analyses were conducted across demographic (age at sample draw), lifestyle (BMI, smoking status, cortisol levels), and social (income, education, insurance status, recent healthcare access) contexts. Age at sample draw was grouped into three categories: ≤55 years, 55–70 years, and >70 years. We repeated age-stratified analyses using age at diagnosis for cases and current age for controls, similar to previous investigations^2–4^. Age at diagnosis was defined as the age at the condition start datetime, reflecting the earliest documented onset of prostate cancer–related conditions. Since age at diagnosis was unavailable for some participants, this analysis was limited to 7,058 cases (n=519 excluded). Further, this approach may introduce bias or residual confounding from not properly accounting for age differences between cases and controls. BMI categories were defined using ancestry-specific thresholds: for non-Asian participants, underweight (<18.5 kg/m²), normal weight (18.5– 24.9 kg/m²), overweight (25.0–29.9 kg/m²), and obesity (≥30.0 kg/m²); for Asian participants, including those classified as SAS or EAS ancestry, underweight (<18.5 kg/m²), normal weight (18.5–22.9 kg/m²), overweight (23.0–24.9 kg/m²), and obesity (≥25 kg/m²)^36^. Smoking status was classified as never, former, or current smoker, based on self-reported survey responses. Morning serum cortisol levels were categorized into three groups according to defined thresholds: low (0–6 μg/dL), normal (6–18 μg/dL), and high (>18 μg/dL). Income was categorized into low (annual income less than 35k), middle (35k-100k), and high (more than 100k) income. Educational attainment was categorized into less than high school, high school or some college, and college graduate or more. Health insurance status was a dichotomized variable indicated whether the participant had health insurance based on self-reported survey responses. Recent healthcare access was determined according to whether participants had seen or spoken to a healthcare provider within the past year based on self-reported survey responses.

### Genetic Data

Whole genome sequencing (WGS) data were obtained from the AoU Research Program, version 8 release. Analyses used the short-read WGS (srWGS) SNP and indel dataset from the ACAF-filtered smaller callset, which includes variants meeting a population-specific allele frequency (AF) greater than 1% or a population-specific allele count (AC) greater than 100 in at least one computed ancestry subpopulation. Variants failing these thresholds, failing quality control filters with a non-PASS flag, or not included in targeted regions were excluded from this callset. This investigation used the previously reported multi-ancestry prostate cancer PRS of 451 variants derived from a large GWAS meta-analysis including 122,188/604,640 (cases/controls) of European ancestry, 19,391/61,608 of African ancestry, 10,809/95,790 of East Asian ancestry, and 3,931/26,405 of Hispanic ethnicity^2^. The variants were extracted from the multi-allelic split Hail MatrixTable and converted to PLINK format for analysis, with 446 variants present in AoU for PRS construction.

When selecting variants, it is possible to retrieve multiple entries corresponding to the same locus, including instances where the reference and alternate alleles of indels are reversed (“flipped”). While not all indels exhibit this pattern, a subset appears in both orientations, with allele frequencies that do not match and do not reflect the flipped alleles. In these instances of duplicate indels retrieved, we compared alleles frequencies to the previously reported PRS allele frequencies, and the orientation matching the intended PRS variant frequency was retained. The PRS was then calculated for each participant as a weighted sum of the number of risk alleles carried, weighting each variant using the previously reported multi-ancestry variant-specific conditional weights^2^.

### Statistical Analyses

The PRS was evaluated as a continuous variable standardized by the mean and standard deviation within each population. Associations between the PRS and prostate cancer status were evaluated using logistic regression models including covariates for age at sample draw and the first 10 principal components of ancestry. Effect sizes were reported as odds ratios per standard deviation unit increase in PRS, along with 95% confidence intervals derived from the model estimates. PRS were also evaluated using an indicator variable splitting the PRS into the following categories: [0-10%], (10-20%], (20-30%], (30-40%], (40-60%], (60-70%], (70-80%], (80-90%], and (90-100%]. Logistic regression models were used to calculate the prostate cancer odds ratios for each PRS category, with the 40–60% PRS category serving as the reference for categorical PRS analyses. Statistical significance was defined as an unadjusted P<0.05.

Area under the curve (AUC) was used to assess the discriminative ability of the PRS. AUCs were calculated using logistic regression for 1) a base model with age at sample draw and top ten principal components of ancestry and 2) a full model with age, principal components, and the continuous PRS. The DeLong test was applied to compare whether the AUCs of the two models significantly differed.

Context-dependent effects of the PRS were evaluated in analyses stratified by demographic, lifestyle, and social contexts. Within each stratum, logistic regression models were used to estimate PRS associations with prostate cancer, adjusting for age and the first 10 principal components. Formal tests of heterogeneity for each context were conducted by comparing PRS effects of each stratum within a context using the Cochrane Q-test. We further evaluated evidence of context-dependent effects in full models (not stratified by contextual factors) that included an interaction term between the continuous PRS and contextual variable, adjusting for the main effects of each. Likelihood ratio tests that compared models with and without the interaction term were also evaluated. Results for all three evaluations are provided in the supplement, with results in the figures and presented in the manuscript text based on heterogeneity P-values from Cochrane Q-test. Statistical significance was defined as an unadjusted P<0.05.

To evaluate the broader clinical relevance of the PRS, a PheWAS was conducted. Using the PheWAS R package^37^, we conducted logistic regression analyses to examine associations between the standardized PRS and phenotypes in male sex participants and separately across sexes to determine whether the PRS has implications for conditions that are not male-specific. Models were adjusted for age at sample collection and the first ten principal components, with sex additionally included as a covariate in analyses across sexes. International Classification of Diseases (ICD) codes were mapped to phecodes using version 1.2 for ICD-9-CM and the 2018 beta version for ICD-10-CM. Analyses were restricted to phecodes with at least 20 cases. Statistical significance was defined as a Bonferroni correction based on the number of phecodes with non-missing p-values, corresponding to p<0.05/1628 = 3.07×10^-5^ in the male-only PheWAS and p<0.05/1813 = 2.76×10^-5^ in the PheWAS across sexes.

---

**Supplemental Figure 1.**
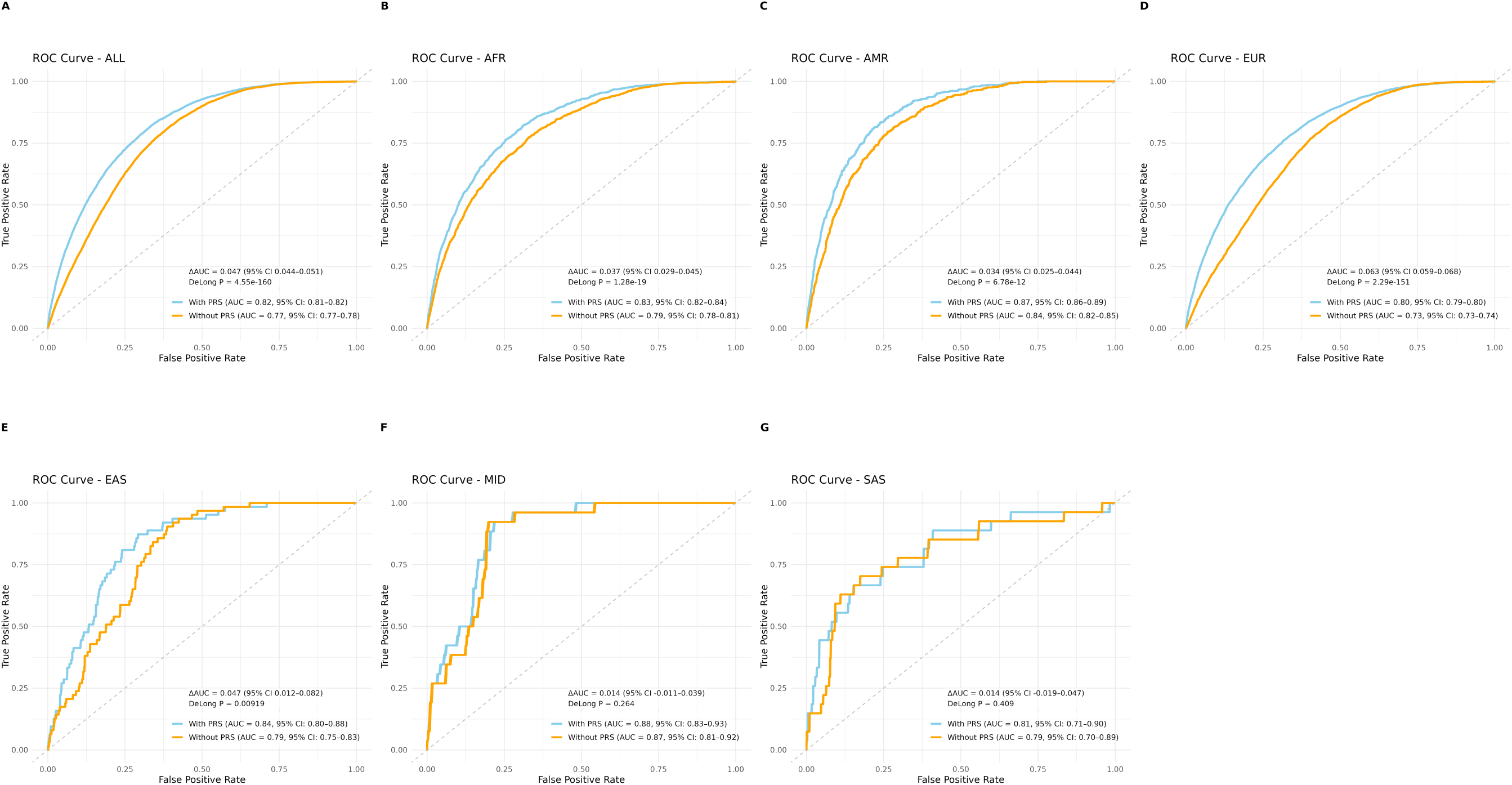
ROC curves of prostate cancer model discrimination with (blue) and without (orange) PRS in (A) All populations and six genetic ancestry groups: (B) AFR, (C) AMR, (D) EUR, (E) EAS, (F) MID, and (G) SAS.

**Supplemental Figure 2.**
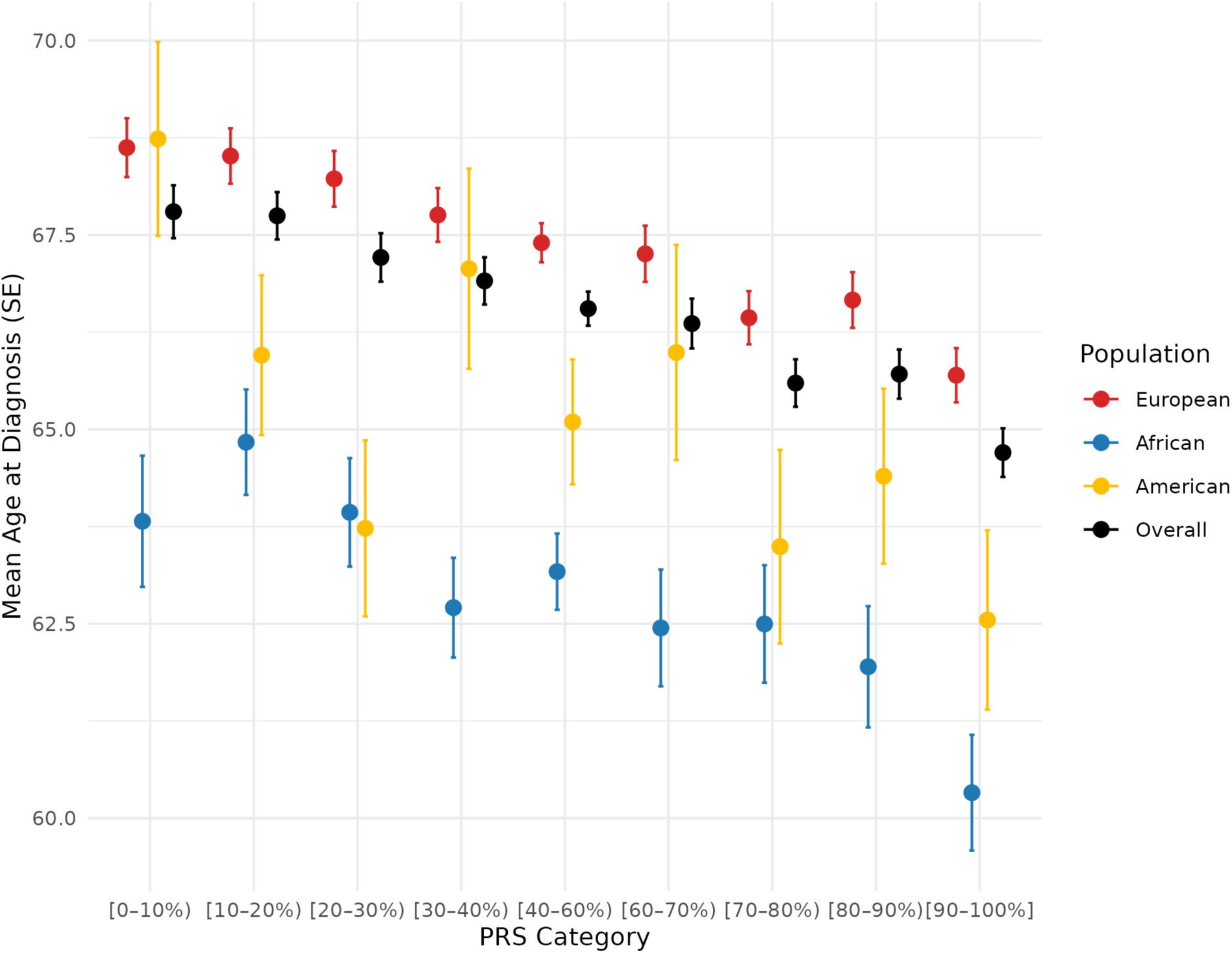
Mean age (years) at prostate cancer diagnosis across PRS categories, stratified by genetic ancestry.

Supplemental Table 1. PRS associations with prostate cancer risk stratified by genetic ancestry groups in AoU. Note that results for PRS deciles are limited to AFR, AMR, and EUR ancestry populations due to the limited number of cases in other ancestry populations.

Supplemental Table 2. Context-dependent associations between PRS and prostate cancer risk.

Supplemental Table 3. Mean age at prostate cancer diagnosis by PRS deciles.

Supplemental Table 4. Prostate cancer PRS PheWAS results in male sex participants.

Supplemental Table 5. Prostate cancer PRS PheWAS results across sexes.

## Notes

### Competing Interest Statement

The authors have declared no competing interest.

### Funding Statement

This study was supported by the National Cancer Institute at the National Institutes of Health (R00 CA246063, R03 CA287235, U01 CA261339, Pacific Northwest Prostate Cancer SPORE P50 CA097186, Cancer Center Support Grant P30 CA015704), The Fred Hutch Translational Data Science New Collaboration Award, an award from the Andy Hill Cancer Research Endowment Distinguished Researchers Program, a Fred Hutch/University of Washington SPORE Career Enhancement Program award, the Prostate Cancer Foundation (21YOUN11), and the Institute for Prostate Cancer Research.

### Author Declarations

This study used data from the All of Us Research Program's Controlled Tier Dataset version 8, available to authorized users on the Researcher Workbench

## References

1. Cancer Statistics Review, 1975-2017 - SEER Statistics https://seer.cancer.gov/archive/csr/1975_2017/index.html.

2. Wang, A., Shen, J., Rodriguez, A.A., Saunders, E.J., Chen, F., Janivara, R., Darst, B.F., Sheng, X., Xu, Y., Chou, A.J., et al. (2023). Characterizing prostate cancer risk through multi-ancestry genome-wide discovery of 187 novel risk variants. Nat Genet 55, 2065–2074. 10.1038/S41588-023-01534-4.

3. Conti, D. V., Darst, B.F., Moss, L.C., Saunders, E.J., Sheng, X., Chou, A., Schumacher, F.R., Olama, A.A. Al, Benlloch, S., Dadaev, T., et al. (2021). Trans-ancestry genome-wide association meta-analysis of prostate cancer identifies new susceptibility loci and informs genetic risk prediction. Nature Genetics 2021 53:1 *53*, 65–75. 10.1038/s41588-020-00748-0.

4. Chen, F., Darst, B.F., Madduri, R.K., Rodriguez, A.A., Sheng, X., Rentsch, C.T., Andrews, C., Tang, W., Kibel, A.S., Plym, A., et al. (2022). Validation of a Multi-ancestry Polygenic Risk Score and Age-Specific Risks of Prostate Cancer: A Meta-analysis Within Diverse Populations. Elife 11. 10.7554/ELIFE.78304.

5. Darst, B.F., Shen, J., Madduri, R.K., Rodriguez, A.A., Xiao, Y., Sheng, X., Saunders, E.J., Dadaev, T., Brook, M.N., Hoffmann, T.J., et al. (2023). Evaluating approaches for constructing polygenic risk scores for prostate cancer in men of African and European ancestry. Am J Hum Genet 110, 1200–1206. 10.1016/j.ajhg.2023.05.010.

6. Kachuri, L., Chatterjee, N., Hirbo, J., Schaid, D.J., Martin, I., Kullo, I.J., Kenny, E.E., Pasaniuc, B., Auer, P.L., Conomos, M.P., et al. (2023). Principles and methods for transferring polygenic risk scores across global populations. Nat Rev Genet 25, 8. 10.1038/S41576-023-00637-2.

7. Vince, R.A., Jiang, R., Bank, M., Quarles, J., Patel, M., Sun, Y., Hartman, H., Zaorsky, N.G., Jia, A., Shoag, J., et al. (2023). Evaluation of Social Determinants of Health and Prostate Cancer Outcomes Among Black and White Patients: A Systematic Review and Meta-analysis. JAMA Netw Open 6, e2250416–e2250416. 10.1001/JAMANETWORKOPEN.2022.50416.

8. Plym, A., Zhang, Y., Stopsack, K.H., Delcoigne, B., Wiklund, F., Haiman, C., Kenfield, S.A., Kibel, A.S., Giovannucci, E., Penney, K.L., et al. (2023). A Healthy Lifestyle in Men at Increased Genetic Risk for Prostate Cancer. Eur Urol 83, 343–351. 10.1016/j.eururo.2022.05.008.

9. Kenfield, S.A., Batista, J.L., Jahn, J.L., Downer, M.K., Van Blarigan, E.L., Sesso, H.D., Giovannucci, E.L., Stampfer, M.J., and Chan, J.M. (2016). Development and Application of a Lifestyle Score for Prevention of Lethal Prostate Cancer. JNCI: Journal of the National Cancer Institute 108. 10.1093/JNCI/DJV329.

10. Benke, I.N., Leitzmann, M.F., Behrens, G., and Schmid, D. (2018). Physical activity in relation to risk of prostate cancer: A systematic review and meta-analysis. Annals of Oncology 29, 1154–1179. 10.1093/annonc/mdy073.

11. Friedenreich, C.M., Wang, Q., Neilson, H.K., Kopciuk, K.A., McGregor, S.E., and Courneya, K.S. (2016). Physical Activity and Survival After Prostate Cancer. Eur Urol 70, 576–585. 10.1016/j.eururo.2015.12.032.

12. Di Maso, M., Augustin, L.S.A., Toffolutti, F., Stocco, C., Dal Maso, L., Jenkins, D.J.A., Fleshner, N.E., Serraino, D., and Polesel, J. (2021). Adherence to mediterranean diet, physical activity and survival after prostate cancer diagnosis. Nutrients 13, 1–11. 10.3390/NU13010243.

13. Jochems, S.H.J., Fritz, J., Häggström, C., Järvholm, B., Stattin, P., and Stocks, T. (2023). Smoking and Risk of Prostate Cancer and Prostate Cancer Death: A Pooled Study. Eur Urol 83, 422–431. 10.1016/j.eururo.2022.03.033.

14. Hurwitz, L.M., Dogbe, N., Barry, K.H., Koutros, S., and Berndt, S.I. (2023). Obesity and prostate cancer screening, incidence, and mortality in the Prostate, Lung, Colorectal, and Ovarian Cancer Screening Trial. JNCI Journal of the National Cancer Institute 115, 1506. 10.1093/JNCI/DJAD113.

15. Genkinger, J.M., Wu, K., Wang, M., Albanes, D., Black, A., van den Brandt, P.A., Burke, K.A., Cook, M.B., Gapstur, S.M., Giles, G.G., et al. (2020). Measures of body fatness and height in early and mid-to-late adulthood and prostate cancer: risk and mortality in The Pooling Project of Prospective Studies of Diet and Cancer. Annals of Oncology 31, 103–114. 10.1016/J.ANNONC.2019.09.007/ATTACHMENT/E9620D15-D710-48F9-98AD-47978EDA3297/MMC3.DOCX.

16. Dickerman, B.A., Torfadottir, J.E., Valdimarsdottir, U.A., Giovannucci, E., Wilson, K.M., Aspelund, T., Tryggvadottir, L., Sigurdardottir, L.G., Harris, T.B., Launer, L.J., et al. (2019). Body fat distribution on computed tomography imaging and prostate cancer risk and mortality in the AGES-Reykjavik study. Cancer 125, 2877–2885. 10.1002/CNCR.32167.

17. Perez-Cornago, A., Dunneram, Y., Watts, E.L., Key, T.J., and Travis, R.C. (2022). Adiposity and risk of prostate cancer death: a prospective analysis in UK Biobank and meta-analysis of published studies. BMC Med 20. 10.1186/S12916-022-02336-X.

18. The “All of Us” Research Program (2019). New England Journal of Medicine 381, 668– 676. 10.1056/NEJMSR1809937.

19. Bick, A.G., Metcalf, G.A., Mayo, K.R., Lichtenstein, L., Rura, S., Carroll, R.J., Musick, A., Linder, J.E., Jordan, I.K., Nagar, S.D., et al. (2024). Genomic data in the All of Us Research Program. Nature 2024 627:8003 *627*, 340–346. 10.1038/s41586-023-06957-x.

20. Bagnato, C.B., Bianco, A., Bonfiglio, C., Franco, I., Verrelli, N., Carella, N., Shahini, E., Zappimbulso, M., Giannuzzi, V., Pesole, P.L., et al. (2024). Healthy Lifestyle Changes Improve Cortisol Levels and Liver Steatosis in MASLD Patients: Results from a Randomized Clinical Trial. Nutrients 16, 4225. 10.3390/NU16234225/S1.

21. Fukuda, S., and Morimoto, K. (2001). Lifestyle, stress and cortisol response: Review II: Lifestyle. Environ Health Prev Med 6, 15. 10.1007/BF02897304.

22. Gandaglia, G., Abdollah, F., Schiffmann, J., Trudeau, V., Shariat, S.F., Kim, S.P., Perrotte, P., Montorsi, F., Briganti, A., Trinh, Q.D., et al. (2014). Distribution of metastatic sites in patients with prostate cancer: A population-based analysis. Prostate 74, 210–216. 10.1002/PROS.22742.

23. Fritsche, L.G., Gruber, S.B., Wu, Z., Schmidt, E.M., Zawistowski, M., Moser, S.E., Blanc, V.M., Brummett, C.M., Kheterpal, S., Abecasis, G.R., et al. (2018). Association of Polygenic Risk Scores for Multiple Cancers in a Phenome-wide Study: Results from The Michigan Genomics Initiative. Am J Hum Genet 102, 1048. 10.1016/J.AJHG.2018.04.001.

24. Graff, R.E., Cavazos, T.B., Thai, K.K., Kachuri, L., Rashkin, S.R., Hoffman, J.D., Alexeeff, S.E., Blatchins, M., Meyers, T.J., Leong, L., et al. (2021). Cross-cancer evaluation of polygenic risk scores for 16 cancer types in two large cohorts. Nat Commun 12, 970. 10.1038/S41467-021-21288-Z.

25. Plym, A., Penney, K.L., Kalia, S., Kraft, P., Conti, D. V., Haiman, C., Mucci, L.A., and Kibel, A.S. (2022). Evaluation of a Multiethnic Polygenic Risk Score Model for Prostate Cancer. J Natl Cancer Inst 114, 771–774. 10.1093/JNCI/DJAB058.

26. Thomas, M., Sakoda, L.C., Hoffmeister, M., Rosenthal, E.A., Lee, J.K., van Duijnhoven, F.J.B., Platz, E.A., Wu, A.H., Dampier, C.H., de la Chapelle, A., et al. (2020). Genome-wide Modeling of Polygenic Risk Score in Colorectal Cancer Risk. Am J Hum Genet 107, 432–444. 10.1016/j.ajhg.2020.07.006.

27. Marston, N.A., Pirruccello, J.P., Melloni, G.E.M., Koyama, S., Kamanu, F.K., Weng, L.C., Roselli, C., Kamatani, Y., Komuro, I., Aragam, K.G., et al. (2023). Predictive Utility of a Coronary Artery Disease Polygenic Risk Score in Primary Prevention. JAMA Cardiol 8, 130–137. 10.1001/JAMACARDIO.2022.4466.

28. Popovici, D., Stanisav, C., Pricop, M., Dragomir, R., Saftescu, S., and Ciurescu, D. (2023). Associations between Body Mass Index and Prostate Cancer: The Impact on Progression-Free Survival. Medicina (B Aires) 59, 289. 10.3390/MEDICINA59020289.

29. Alavanja, M.C.R., Samanic, C., Dosemeci, M., Lubin, J., Tarone, R., Lynch, C.F., Knott, C., Thomas, K., Hoppin, J.A., Barker, J., et al. (2003). Use of agricultural pesticides and prostate cancer risk in the agricultural health study cohort. Am J Epidemiol 157, 800–814. 10.1093/AJE/KWG040.

30. DeRouen, M.C., Schupp, C.W., Koo, J., Yang, J., Hertz, A., Shariff-Marco, S., Cockburn, M., Nelson, D.O., Ingles, S.A., John, E.M., et al. (2018). Impact of individual and neighborhood factors on disparities in prostate cancer survival. Cancer Epidemiol 53, 1–11. 10.1016/j.canep.2018.01.003.

31. Iyer, H.S., James, P., Valeri, L., Hart, J.E., Pernar, C.H., Mucci, L.A., Holmes, M.D., Laden, F., and Rebbeck, T.R. (2020). The association between neighborhood greenness and incidence of lethal prostate cancer: A prospective cohort study. Environmental Epidemiology 4. 10.1097/EE9.0000000000000091.

32. Szymanski, K.M., Wheeler, D.C., and Mucci, L.A. (2010). Fish consumption and prostate cancer risk: A review and meta-analysis. American Journal of Clinical Nutrition 92, 1223–1233. 10.3945/ajcn.2010.29530.

33. Zu, K., Mucci, L., Rosner, B.A., Clinton, S.K., Loda, M., Stampfer, M.J., and Giovannucci, E. (2014). Dietary lycopene, angiogenesis, and prostate cancer: A prospective study in the prostate-specific antigen era. J Natl Cancer Inst 106. 10.1093/JNCI/DJT430.

34. Lennon, N.J., Kottyan, L.C., Kachulis, C., Abul-Husn, N.S., Arias, J., Belbin, G., Below, J.E., Berndt, S.I., Chung, W.K., Cimino, J.J., et al. (2024). Selection, optimization and validation of ten chronic disease polygenic risk scores for clinical implementation in diverse US populations. Nat Med 30, 480–487. 10.1038/S41591-024-02796-Z.

35. All of Us Genomic Quality Report – User Support https://support.researchallofus.org/hc/en-us/articles/29390274413716-All-of-Us-Genomic-Quality-Report.

36. Nishida, C., Barba, C., Cavalli-Sforza, T., Cutter, J., Deurenberg, P., Darnton-Hill, I., Deurenberg-Yap, M., Gill, T., James, P., Ko, G., et al. (2004). Appropriate body-mass index for Asian populations and its implications for policy and intervention strategies. The Lancet 363, 157–163. 10.1016/S0140-6736(03)15268-3.

37. Carroll, R.J., Bastarache, L., and Denny, J.C. (2014). R PheWAS: Data analysis and plotting tools for phenome-wide association studies in the R environment. Bioinformatics 30, 2375–2376. 10.1093/BIOINFORMATICS/BTU197.

